# Empirical Sample Size Determination for Popular Classification Algorithms in Clinical Research

**DOI:** 10.1101/2024.05.03.24306846

**Authors:** Scott Silvey, Jinze Liu

## Abstract

**Motivation:** The performance of a classification algorithm eventually reaches a point of diminishing returns, where additional sample added does not improve results. Thus, there is a need for determining an optimal sample size that both maximizes performance, while accounting for computational burden or budgetary concerns.

**Methods:** Sixteen large open-source datasets were collected, each containing a binary clinical outcome. Four machine learning algorithms were assessed: XGBoost (XGB), Random Forest (RF), Logistic Regression (LR), and Neural Networks (NN). For each dataset, the cross-validated AUC was calculated at increasing sample sizes, and learning curves were fit. Sample sizes needed to reach the full-dataset AUC minus 2% (or, 0.02) were calculated from the fitted learning curves and compared across the datasets and algorithms. Dataset-level characteristics: minority class proportion, full-dataset AUC, strength/number/type of features, and degree of nonlinearity, were examined. Negative binomial regression models were used to quantify relationships between these characteristics and expected sample sizes within each algorithm. Four multivariable models were constructed which selected the best combination of dataset-specific characteristics that minimized out-of-sample prediction error. Additional models were fitted which allowed for prediction of the expected gap in performance at a given sample size using the same empirical learning curve data.

**Results:** Among the sixteen datasets (full-dataset sample sizes ranging from 70,000-1,000,000), median sample sizes were 9,960 (XGB), 3,404 (RF), 696 (LR), and 12,298 (NN) to reach AUC convergence. For all four algorithms, more balanced classes (multiplier: 0.93-0.96 for 1% increase in minority class proportion) were associated with decreased sample size. Other characteristics varied in importance across algorithms - in general, more features, weaker features, and more complex relationships between the predictors and the response increased expected sample sizes. In multivariable analysis, top selected predictors were minority class proportion, full-dataset AUC, and dataset nonlinearity (XGB and RF). For LR, top predictors were minority class proportion, percentage of strong linear features, and number of features. For NN, top predictors were minority class proportion, percentage of numeric features, and dataset nonlinearity.

**Conclusions:** The sample sizes needed to reach convergence among four popular classification algorithms vary by dataset and method and are associated with dataset-specific characteristics that can be influenced or estimated prior to the start of a research study.

## Introduction

Machine learning (ML) is becoming increasingly popular within the domain of healthcare data analysis and clinical decision making [1]. The lack of a fixed model specification and distributional assumptions allows for these methods to learn complex relationships that are not necessarily linear in nature, such as high-order interactions and polynomial effects. Due to this, most popular machine-learning algorithms require much larger sample sizes than traditional statistical methods [2]. However, exact amounts are not clear, and combined with the fact that there are many different ML algorithms (each containing their own limitations and properties) [3], there is a lack of transparency in selecting an optimal sample size for ML analysis. It is known that, for any given dataset, there is a point where adding additional samples will not increase the performance metrics of the model considerably [4]. Thus, it becomes important to collect enough data to optimize these metrics while also accounting for this performance ceiling and the budgetary or computational concerns that may arise when collecting substantial amounts of unnecessary data.

Another reason for difficulty selecting a proper sample size when applying machine learning is the lack of a true endpoint or common metric of interest. The traditional target for sample size determination methods is the statistical power to detect a certain effect size [5]. In machine learning, since predictive performance rather than parameter estimation is usually of interest, this endpoint becomes unclear. A commonly used metric of predictive performance is prediction accuracy, defined as the proportion of correct classifications made [6]. However, the prediction accuracy is related to the distribution of the outcome; for a rare event, accuracy can be high even with a completely non-informative model [7]. As a result, a fairer performance metric is the area under the receiver operating characteristic curve (AUC), which evaluates model predictions over a range of probability thresholds from 0 to 1 [8]. An AUC of 0.5 implies a completely random prediction, while an AUC of 1.0 indicates perfect classification. A desirable property of AUC is insensitivity to the proportion of cases versus controls in the dataset [9]. While the AUC is commonly used to evaluate the performance of an ML algorithm, other metrics such as the under the precision-recall curve [10] can also be preferred. Thus, the metric of choice for a certain model depends on the clinical question of interest.

### Related Works

The concept of empirically estimating the performance of a classification algorithm as the training set size increases has been widely explored in a variety of different settings. This is typically done by creating a “learning curve,” measuring a metric (such as classification accuracy) as a function of sample size [11]. Perlich et. al [12] compared logistic regression approaches versus decision-tree based approaches, demonstrating that logistic regression often outperforms tree induction in small samples, but decision trees excel as the sample size becomes large. Mukherjee et al [13] developed a method to assess the error rate of a classifier as a function of sample size using an inverse power-law model. Their method was introduced in the context of DNA microarray data, which often contains a large amount of features and limited access to samples due to cost restraints. Figueroa et al. [14] modified the original learning curve fitting process by using nonlinear weighted least squares to favor future predictions, using three moderately sized datasets to demonstrate their algorithm. More recently, Richter and Khoshgoftaar [15] experimented with learning curves on biomedical big data with limited labels and heavy class imbalance, using 1% of the full dataset AUC as their stopping rule. Because the cost of labelling certain types of data is expensive, it is important to maximize the quality of the data while minimizing costs. They found that a semi-supervised approach and pseudo-labelled data generated from a small amount of actual data could accurately predict future performance.

Our current study aims to develop algorithm-specific sample size guidelines using dataset-specific variables that can be estimated or manipulated by researchers before any data has been collected, analogous to a sample size calculation performed in a traditional power analysis. We examined four popular binary classification algorithms in the context of clinical research, where the aim is to predict a health-related outcome such as a disease state or event. Previous contributions have focused on the methodology of learning curve fitting or estimating future performance from an already-collected sample. Previous studies have also mostly used small datasets in the context of -omics type data. In general, a focused clinical study often contains fewer features and fewer correlated features than -omics data [16]. The contributions of this study include a learning curve analysis of sixteen datasets, concrete sample size guidelines based on dataset-specific characteristics, as well as creation of an RShiny app where the guidelines developed from this analysis can be applied.

## Methods

### Dataset Description

We have collected sixteen public-access clinical datasets ranging from sample sizes of 70,000-1,000,000. All datasets contained a single binary outcome, such as a disease state, with a combination of continuous numeric, discrete numeric, or binary predictors. Continuous numeric features were considered to have at least 10 unique values. Dataset characteristics, as well as their source location, were summarized. It should be noted that eight of these sixteen datasets were artificially created from smaller real-life datasets using Bayesian Network Generation, and their details have been previously discussed [17].

A detailed description of specific data pre-processing steps can be found in the supplementary section. In summary, nominal variables were converted to binary variables based on arbitrary binning rules, and variables containing text values (i.e. “gender”: male vs. female) were also converted to binary variables. Missing data was present in three datasets only (CDC Heart Disease (2022), Diabetes130, and COVID-19), although the amount of missingness was quite low among these sets (1.3%, <1.0%, and <1.0%, respectively). Without knowing additional information regarding the nature of these missing values, we considered them missing completely at random (MCAR) and performed mean-imputation [18].

### Learning Curve Approach

From the sixteen datasets studied, we evaluated the cross-validated AUC (CV-AUC) as a function of increasing sample size. Our approach to generate learning curves for each dataset (and each algorithm) was as follows:

- Create a list of proposed training set sizes.
- At each point in the sample size interval, generate ten random samples of size *n* from the full dataset.
- In each of the ten samples, estimate the (five-fold, outcome-stratified) CV-AUC on the proposed algorithm of choice. Average the ten CV-AUC values to generate an estimate of out-of-sample performance at a given *n*.
- Repeat at the next *n* in the list.

For step 1.), the training set size list usually consisted of 10 evenly spaced points ranging from *n* = 500 to *n* = 50,000, but if convergence was not reached by *n* = 50,000, the end point was extended. For logistic regression, the final *n* was lower, as these models typically converged much earlier than more complex ML algorithms. A full description of the sample size intervals used for each dataset and each algorithm can be found in the Supplementary section.

Convergence was defined as the smallest *n* where the CV-AUC was within two percentage points of the full-dataset AUC. For example, if the full-dataset AUC was 0.85, we would obtain the smallest *n* where a CV-AUC of 0.83 was first surpassed. We chose this stopping point (2% from full-dataset AUC) because we believed that it provided the most reasonable trade-off between high performance, computational burden, and sample size. The full-dataset AUC for each classification algorithm was calculated using five-fold stratified cross-validation [19] on the entire dataset.

Once the raw data was generated, estimated learning curves were fit using nonlinear least squares optimization [20], following the power law equation: *AUC_(n)_* = *an^b^* + *c*, where *a* and *b* were estimated, and *c* was either fixed to be the full-dataset AUC or was also estimated, depending on the quality of the fit. For some datasets and algorithms, the power law function did not fit the data well. These were typically scenarios where the dataset required a relatively larger sample size to converge. In these scenarios, we instead fit the learning curves using a logarithmic function, *AUC_(n)_* = β*_0_* + β*_1_*\**log*(*n)*, where β*_0_* and β*_1_* were estimated using ordinary least-squares [21].

### Classification Algorithms

We examined the following binary classifiers on each dataset: logistic regression (LR) [22], random forest (RF) [23], XGBoost (XGB) [24], and neural networks (NN) [25]. These algorithms were selected due to their widespread and popular use in clinical data analysis. We performed logistic regression by fitting a multivariable model using all predictors without any variable selection methods. For the random forest and XGBoost algorithms, hyperparameters were left at their default values [23, 24]. For neural networks, we used the R package h2o [26] to perform our analyses; we considered one hidden layer with 20 units, and 10 epochs of data training.

### Sample Size Determination and Guidelines

Following the learning curve analysis of the four selected algorithms on our datasets, we examined the effects of six dataset-level factors on the predicted sample sizes needed for convergence. These included minority class proportion (maximum value of 50%, indicating no class imbalance), separability (defined as the full-dataset AUC itself), the total number of features, the percentage of features that were continuous (versus binary or discrete numeric), the percentage of “core linear” features, and “dataset nonlinearity.” Core linear features were determined by adding an L1 (LASSO) penalty to the logistic regression model for each full dataset [27]. The percentage of variables that did not shrink to zero when this penalty was added were defined as core linear features. Dataset nonlinearity was a rough measure of the degree of nonlinear or interactive relationships between the predictors and the outcome that were present in the data. This was defined as the percentage point difference in the full-dataset AUC when using a complex algorithm (XGBoost) compared to logistic regression. For example, if logistic regression yielded a full-dataset AUC of 0.90 and XGBoost yielded a full-dataset AUC of 0.95, the dataset nonlinearity would be calculated as 5.0%. For the purpose of calculating these values, XGBoost hyperparameters were left at their default values [24].

Within the context of each algorithm, the relationship between these dataset-specific variables and the *n* required for AUC convergence was examined. Because estimated sample sizes were discrete and right-skewed numeric values, we used negative binomial regression models [28] to quantify the strength and significance of each dataset characteristic on predicted sample sizes, which produce coefficients in terms of log-expected counts. Then, in multivariable negative binomial regression models for each algorithm, we selected up to three dataset-level predictors that together minimized the Akaike Information Criterion, which evaluates how well the model fits the data while penalizing for the number of parameters estimated [29]. A maximum of three predictors per model were considered in order to avoid potential overfitting. We also calculated adjusted deviance-based pseudo-R^2^ statistics [30,31], which further quantified each model’s goodness-of-fit and proportion of deviance explained by the predictors. The final model equations were reported and discussed for each algorithm, and visualizations of the model predictions at varying levels of each dataset-specific characteristic were generated. Statistical significance was set to α = 0.05 for all hypothesis tests considered, and RStudio version 4.2.3 was used for all analyses.

### Expected Performance at Pre-Specified Sample Size

As a secondary endpoint, we explored prediction of the expected difference between the training set AUC at size *n* and the full-dataset AUC. For this, we built a second set of models that used the empirical learning curve data for each of the sixteen datasets collected. The outcome was the number of percentage points away from the full-dataset AUC that was achieved at each sample size point in the learning curve procedure. For example, if a dataset’s AUC was 0.80 and at *n* = 500, an AUC of 0.75 was achieved, the difference calculated at this point would be 5%.

Additionally, this outcome was natural log-transformed. Linear mixed effects models were fitted for each of the four algorithms with a dataset-level random intercept term, and sample size plus the (previously determined) top three dataset-specific characteristics for each algorithm as fixed effects. We also included interaction terms between sample size and each dataset specific characteristic, which modeled the potential difference in slopes across different levels of the covariates. After the models were fitted, we plugged in the expected sample sizes from the primary analysis (which represented an AUC difference of 2%, by definition), and examined the agreement between the output and the true values. Finally, we discussed the implications of using these models in conjunction with the main set of negative binomial models from the primary analysis.

## Results

We assembled sixteen datasets with sample sizes ranging from 70,000 to 1,000,000. Of the four classification algorithms examined, XGB performed the best or tied for the best performance on 14/16 (87.5%) datasets, while random forest performed the best on two. Full dataset AUCs (separability) ranged from 0.608-0.979 (XGB), 0.609-0.976 (RF), 0.596-0.949 (LR), and 0.603-0.974 (NN) (Table 2). As expected, logistic regression models generally performed the worst, with full-dataset AUCs that were 2.8 percentage points lower on average compared to XGB.

### Learning Curve Results

Learning curves were fit to the sixteen collected datasets. Table 2 and Figure 1 contain a full summary and visualization of estimated sample sizes across each classification algorithm and dataset. Neural networks required the largest sample sizes to reach convergence, and also had the most variability among the datasets (median *n* = 12,298, range: 1,824-180,835). Logistic regression required the smallest sample size to converge, and also was the least variable (median *n* : 696, range : 204-6,798). XGB required approximately three times the sample size compared to RF, but the range of estimated sample sizes generated from RF models was nearly twice as wide (Table 2). Figure 2 shows the fitted learning curves for each algorithm generated within each dataset, with a marker indicating the earliest sample size where the CV-AUC was within two percentage points of the full-dataset AUC.

**Figure 1:**
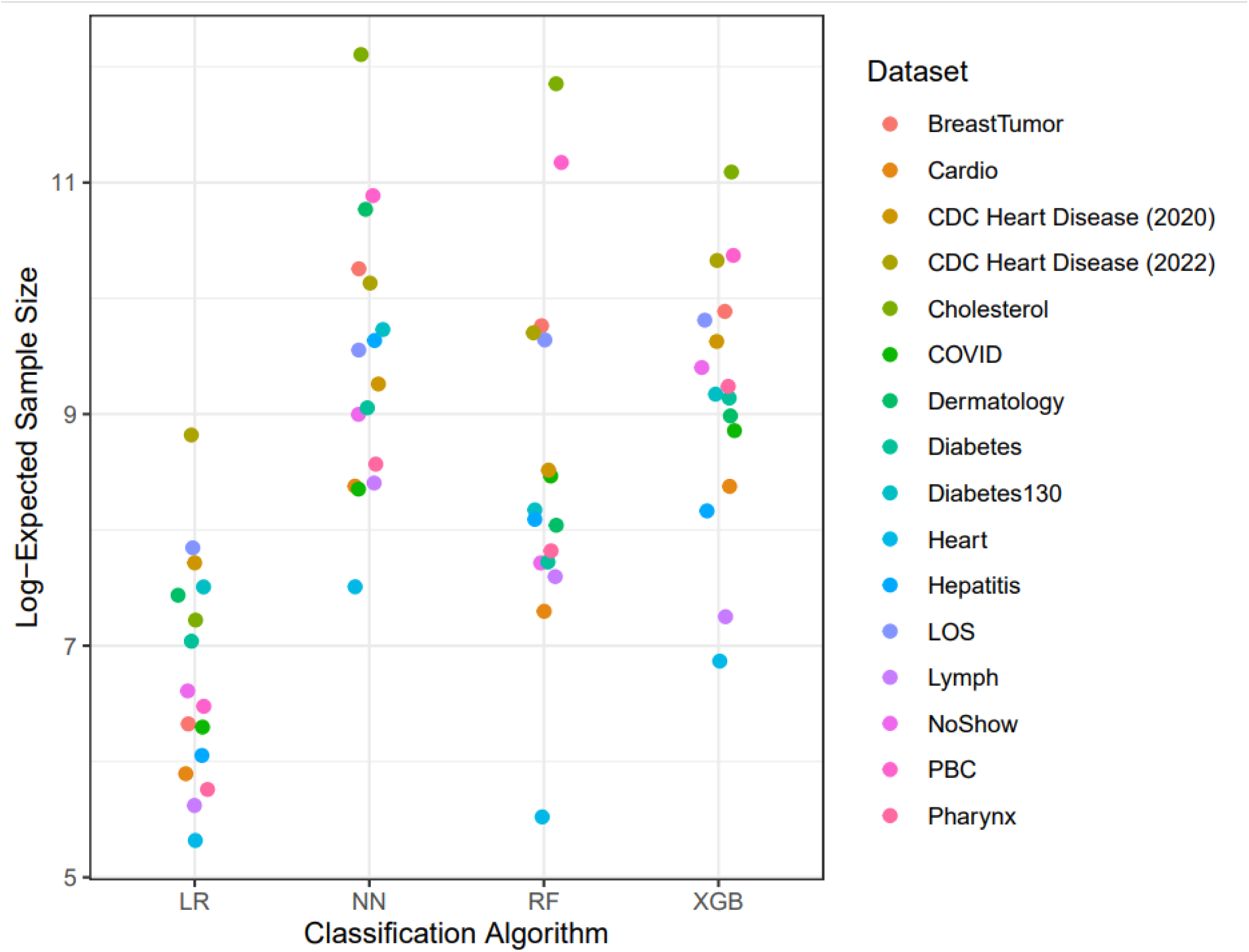
Visualization of expected sample sizes calculated from the learning-curve analysis of sixteen datasets. The y-axis represents the natural-log transformed sample size values, while the x-axis is grouped by classification algorithm. Each dataset is represented by a different color.

**Figure 2:**
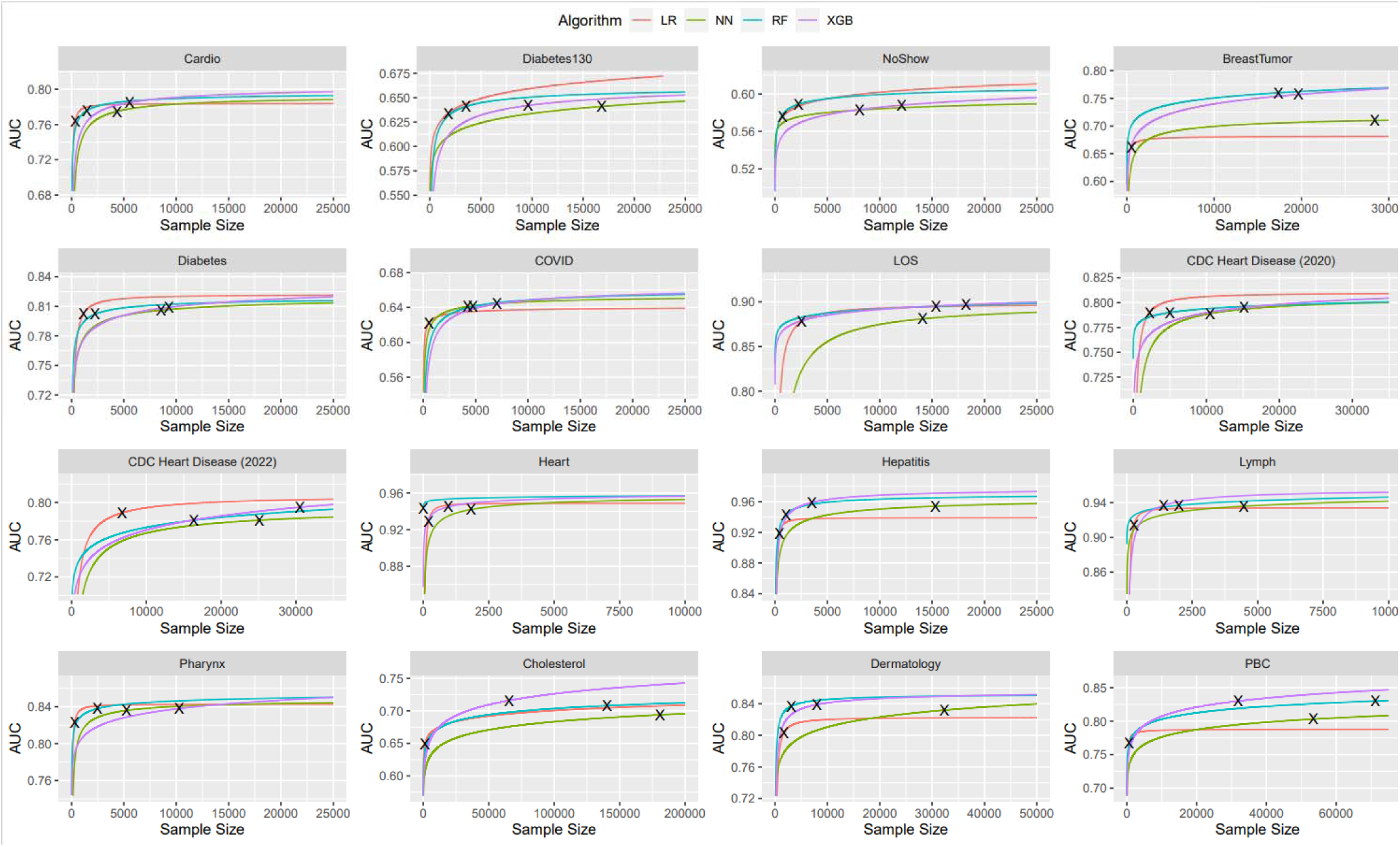
Fitted learning curves for sixteen datasets across four classification algorithms. The x-axis is sample size, while the y-axis is area-under the receiver operating characteristic curve (AUC). Different colors were chosen for different algorithms. Each black X represents the point where the AUC at size *n* first comes within 2% (or, 0.02) of the asymptotic (i.e. full-dataset) AUC.

### Dataset Specific Characteristics

Dataset-specific characteristics were examined (Table 1). The average minority class percentage was 25.18% (± 15.93%) and average number of features was 18 (± 9). The average percentage of continuous numeric features was 24.60% (± 15.31%), and the average percentage of core linear features was 67.69% (± 23.73%). The median dataset nonlinearity was 1.85% (range: 0.50-9.50%). Most datasets (13/16, 81.3%) had nonlinearity values under 5.0%. Thus, for the purpose of model fitting, this was converted into a binary variable indicating either high (≥ 5.0%) or low (< 5.0%). Scatterplots examining the visual relationships between log-expected sample sizes and each dataset-specific characteristic can be found in Figure 3.

**Table 1:** Dataset-Specific Characteristics

| Dataset Name | Source | Full Dataset Size | Minority Class Proportion | Features | Core Linear Features (%) | Continuous Features (%) | Nonlinearity |
| --- | --- | --- | --- | --- | --- | --- | --- |
| Cardio | OpenML | 70,000 | 50.0% | 11 | 100.0% | 45.5% | 1.8% |
| Diabetes130 | OpenML | 101,766 | 46.1% | 35 | 45.7% | 20.0% | 0.8% |
| NoShow | OpenML | 110,527 | 20.2% | 8 | 25.0% | 12.5% | 1.2% |
| BreastTumor | OpenML | 116,640 | 34.6% | 9 | 44.4% | 33.3% | 9.5% |
| Diabetes | UCI | 253,680 | 13.9% | 21 | 85.7% | 19.0% | 0.7% |
| COVID-19 | OpenML | 263,007 | 39.0% | 16 | 37.5% | 6.25% | 2.2% |
| LOS | OpenML | 318,438 | 2.1% | 11 | 36.4% | 27.3% | 1.9% |
| CDC Heart Disease (2020) | Kaggle | 319,795 | 4.4% | 17 | 76.5% | 23.5% | 0.5% |
| CDC Heart Disease (2022) | Kaggle | 394,509 | 8.6% | 39 | 59.0% | 15.4% | 0.6% |
| Heart | OpenML | 1,000,000 | 44.4% | 13 | 92.3% | 46.2% | 1.6% |
| Hepatitis | OpenML | 1,000,000 | 20.8% | 19 | 94.7% | 31.6% | 4.0% |
| Lymph | OpenML | 1,000,000 | 45.7% | 18 | 83.3% | 5.6% | 2.3% |
| Pharynx | OpenML | 1,000,000 | 25.6% | 11 | 72.2% | 18.2% | 1.5% |
| Cholesterol | OpenML | 1,000,000 | 16.5% | 13 | 61.5% | 30.8% | 6.7% |
| Dermatology | OpenML | 1,000,000 | 13.2% | 33 | 84.8% | 3.0% | 3.6% |
| PBC | OpenML | 1,000,000 | 17.8% | 18 | 83.3% | 55.6% | 6.2% |

**Table 2:**
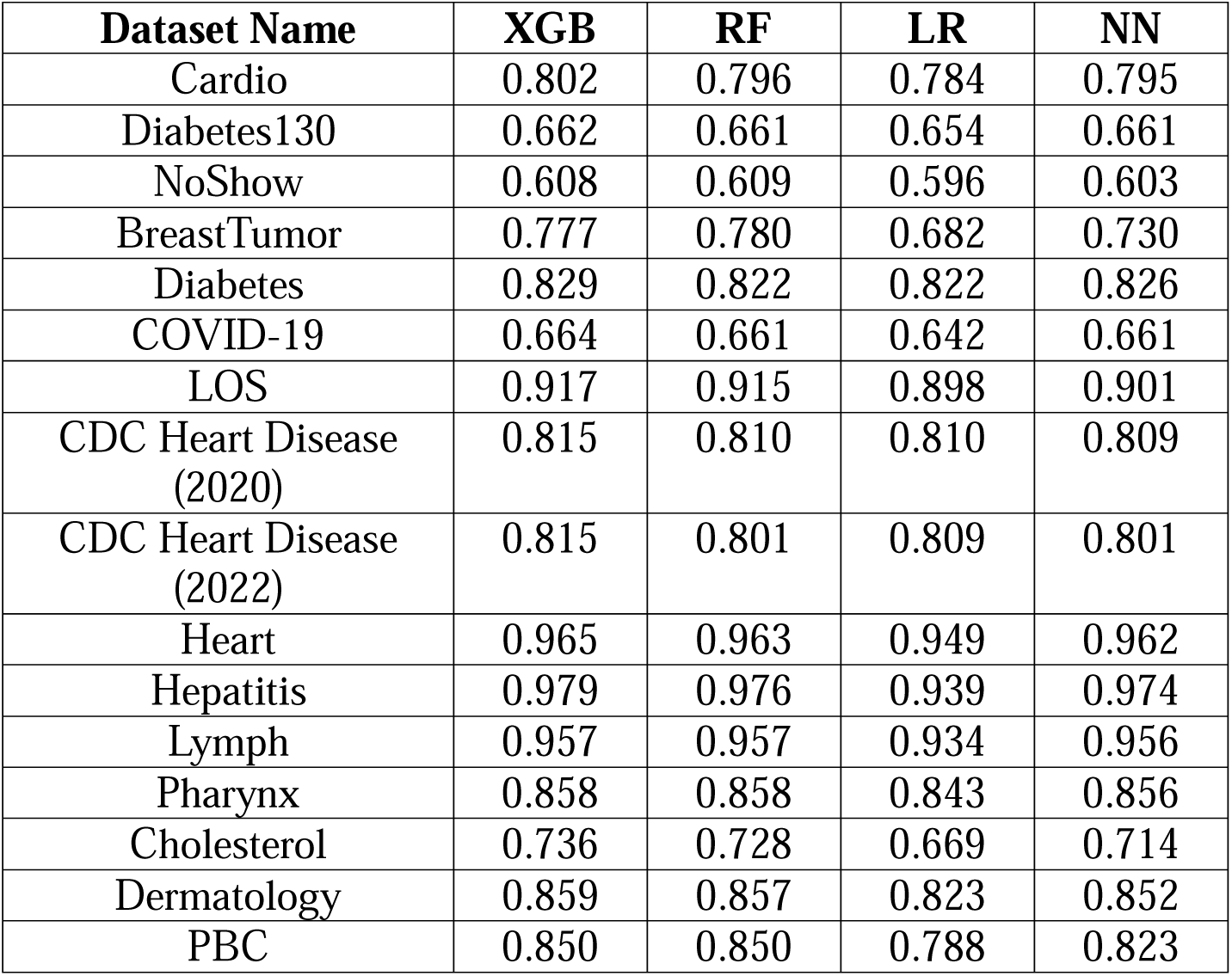
Full-Dataset AUC (Separability) for Each Algorithm.

| **Dataset Name** | **XGB** | **RF** | **LR** | **NN** |
| --- | --- | --- | --- | --- |
| Cardio | 0.802 | 0.796 | 0.784 | 0.795 |
| Diabetes130 | 0.662 | 0.661 | 0.654 | 0.661 |
| NoShow | 0.608 | 0.609 | 0.596 | 0.603 |
| BreastTumor | 0.777 | 0.780 | 0.682 | 0.730 |
| Diabetes | 0.829 | 0.822 | 0.822 | 0.826 |
| COVID-19 | 0.664 | 0.661 | 0.642 | 0.661 |
| LOS | 0.917 | 0.915 | 0.898 | 0.901 |
| CDC Heart Disease<br>(2020) | 0.815 | 0.810 | 0.810 | 0.809 |
| CDC Heart Disease<br>(2022) | 0.815 | 0.801 | 0.809 | 0.801 |
| Heart | 0.965 | 0.963 | 0.949 | 0.962 |
| Hepatitis | 0.979 | 0.976 | 0.939 | 0.974 |
| Lymph | 0.957 | 0.957 | 0.934 | 0.956 |
| Pharynx | 0.858 | 0.858 | 0.843 | 0.856 |
| Cholesterol | 0.736 | 0.728 | 0.669 | 0.714 |
| Dermatology | 0.859 | 0.857 | 0.823 | 0.852 |
| PBC | 0.850 | 0.850 | 0.788 | 0.823 |

**Figure 3:**
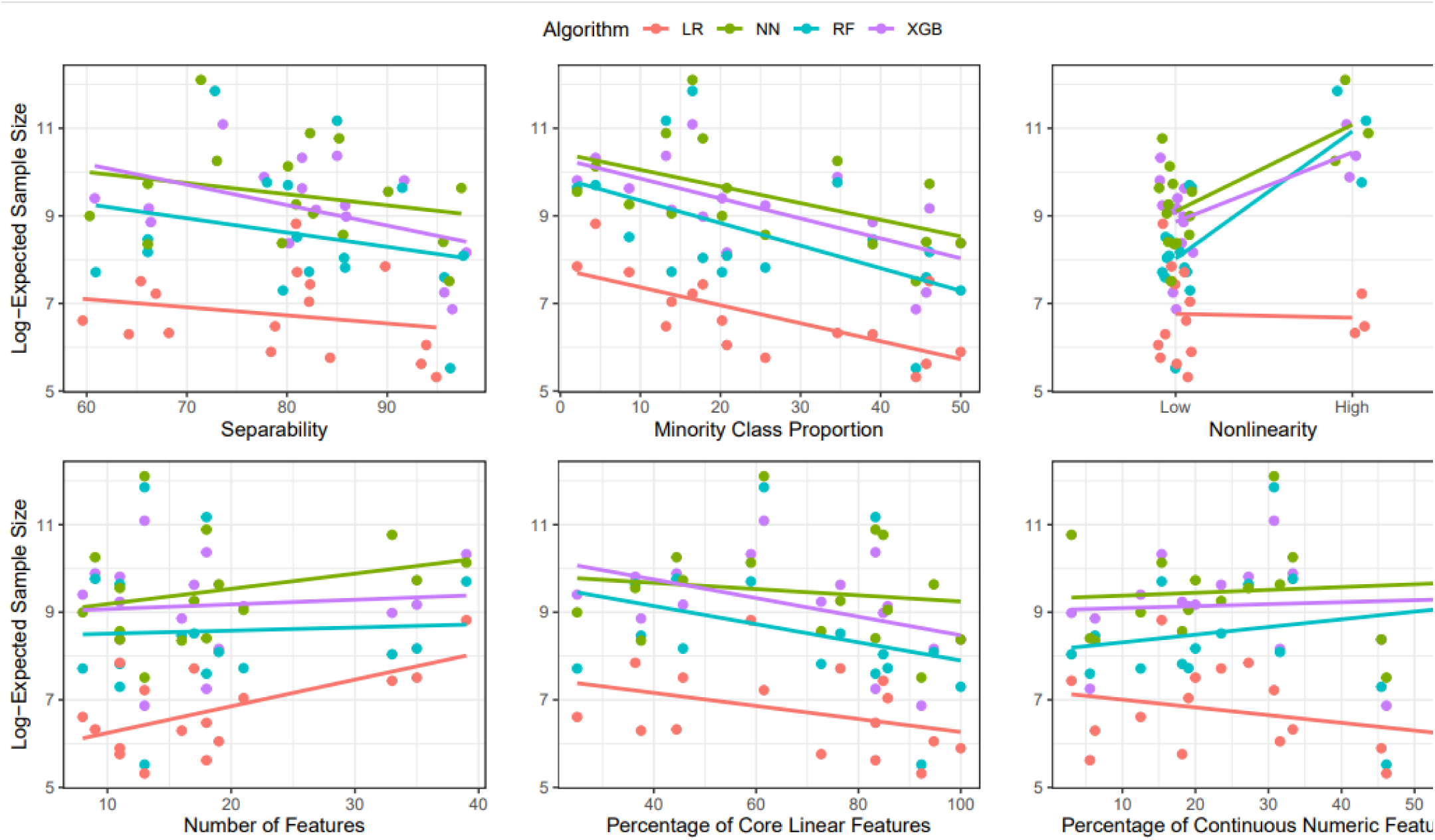
Relationships between each dataset-specific characteristic and expected sample sizes. The y-axis represents the natural-log transformed sample size values, while each x-axis represents varying levels of each dataset-specific characteristic. Different colors were chosen for different algorithms. Lines represent simple lines-of-best fit, included for the purpose of visualization. Separability multiplied by 100. All values representing proportions were multiplied by 100 so that 0 indicates 0% and 100 indicates 100%. Nonlinearity “low” : < 5%, “high” : ≥ 5%.

Negative binomial regression models were fitted, examining the individual associations between each of the dataset-specific characteristics and predicted sample sizes (Table 3). In these models, separability (full-dataset AUC) was multiplied by 100 for easier interpretation. For example, an AUC of 0.80 was entered as 80.0 in the models. For XGBoost, minority class proportion and separability were both inversely related with sample size; for every 1% increase in separability (where 50.0 was the baseline value), estimated sample sizes were affected by a multiplier of 0.955 (*p*=0.024). For every 1% increase in minority class proportion, estimated samples sizes were affected by a multiplier of 0.959 (*p*=0.0007). In datasets with high (≥ 5.0%) values of nonlinearity, estimated sample sizes were affected by a multiplier of 3.888 (*p*=0.005). In the Random Forest analyses, results were similar for minority class proportion (0.931× multiplier for 1% increase, *p*=0.0003), separability (0.939× multiplier for each 1% increase over 50.0, *p*=0.047), and nonlinearity (15.984× multiplier for those with high values, *p*<0.0001). However, the percentage of continuous numeric features (1.065× multiplier for every 1% increase, *p*=0.003) was also individually statistically significant. For logistic regression, minority class proportion (0.963× multiplier for every 1% increase, *p*=0.001), the number of features (1.056× multiplier for each additional feature, *p*=0.006), the percentage of core features (0.982× multiplier for 1% increase, *p*=0.046), and the percentage of continuous numeric features (0.971× multiplier for 1% increase, *p*=0.042) were significantly associated with sample size. Again, a more balanced ratio of classes reduced the needed sample size, while more features increased sample size. However, a higher percentage of core linear features and a higher percentage of continuous numeric features lowered the sample size. Finally, for neural networks, results were similar to XGBoost; minority class proportion (0.953× multiplier for 1% increase, *p*=0.003), full-dataset AUC (0.950× multiplier for each 1% increase over 50.0, *p*=0.031), and nonlinearity (6.85× multiplier for high values, *p*=0.0001) were all individually statistically significant.

**Table 3:** Predicted Sample Sizes Needed to Reach 2% of Full-Dataset AUC from Learning Curve Analysis

| **Dataset Name** | **XGB** | **RF** | **LR** | **NN** |
| --- | --- | --- | --- | --- |
| Cardio | 4,341 | 1,476 | 363 | 4,349 |
| Diabetes130 | 9,623 | 3,544 | 1,822 | 16,823 |
| NoShow | 12,114 | 2,241 | 742 | 8,084 |
| BreastTumor | 19,668 | 17,383 | 558 | 28,424 |
| Diabetes | 9,306 | 2,261 | 1,140 | 8,556 |
| COVID-19 | 7,026 | 4,750 | 543 | 4,241 |
| LOS | 18,239 | 15,381 | 2,555 | 14,085 |
| CDC Heart Disease (2020) | 15,177 | 4,995 | 2,243 | 10,510 |
| CDC Heart Disease (2022) | 30,534 | 16,355 | 6,768 | 25,120 |
| Heart | 960 | 250 | 204 | 1,824 |
| Hepatitis | 3,513 | 3,265 | 425 | 15,302 |
| Lymph | 1,409 | 1,992 | 276 | 4,470 |
| Pharynx | 10,296 | 2,488 | 317 | 5,260 |
| Cholesterol | 65,556 | 140,499 | 1,368 | 180,835 |
| Dermatology | 7,979 | 3,103 | 1,696 | 47,489 |
| PBC | 31,897 | 71,194 | 650 | 53,453 |
| **Median (Range)** | 9,960 (960 – 65,556) | 3,404 (250-140,499) | 696 (204-6,798) | 12,298 (1824-180,835) |
| **Mean (SD), Log-Transform** | 9.16 (1.11) | 8.57 (1.55) | 6.75 (0.96) | 9.47 (1.17) |

**Table 4:** Univariable Association of Each Dataset-Specific Characteristic with Predicted Sample Size

| Variable | XGB | RF | LR | NN |
| --- | --- | --- | --- | --- |
| Minority Class Proportion | 0.959 [0.934-0.985], ***p*=0.0007** | 0.931 [0.891-0.977], ***p*=0.0003** | 0.963 [0.945-0.983], ***p*=0.001** | 0.953 [0.921-0.990], ***p*=0.003** |
| Separability | 0.955 [0.903-1.000], ***p*=0.024** | 0.939 [0.861-1.028], ***p*=0.047** | 0.999 [0.946-1.051], *p*=0.976 | 0.950 [0.898-1.007], ***p*=0.031** |
| Number of Features | 1.000 [0.958-1.053], *p*=0.995 | 0.966 [0.904-1.059], *p*=0.354 | 1.056 [1.021-1.097], ***p*=0.006** | 1.001 [0.947-1.071], *p*=0.981 |
| Continuous Features (%) | 1.019 [0.986-1.057], *p*=0.211 | 1.065 [1.013-1.121], ***p*=0.003** | 0.971 [0.942-1.006], ***p*=0.042** | 1.019 [0.982-1.059], *p*=0.04 |
| Core Linear Features (%) | 0.983 [0.958-1.001], *p*=0.071 | 0.988 [0.941-1.034], *p*=0.413 | 0.982 [0.959-1.004], ***p*=0.046** | 0.994 [0.960-1.028], *p*=0.610 |
| Dataset Nonlinearity | 3.889 [1.631-11.209], ***p*=0.005** | 15.985 [5.914-55.754], ***p*<0.0001** | 0.585 [0.211-2.112], *p*=0.346 | 6.853 [2.763-20.947], ***p*=0.0001** |

In multivariable models for each algorithm, we selected the set of three predictors that minimized the AIC. The equation below, as well as Table 5, presents a summary of each algorithm-specific model, which shows the adjusted contribution of each predictor to the expected sample size.

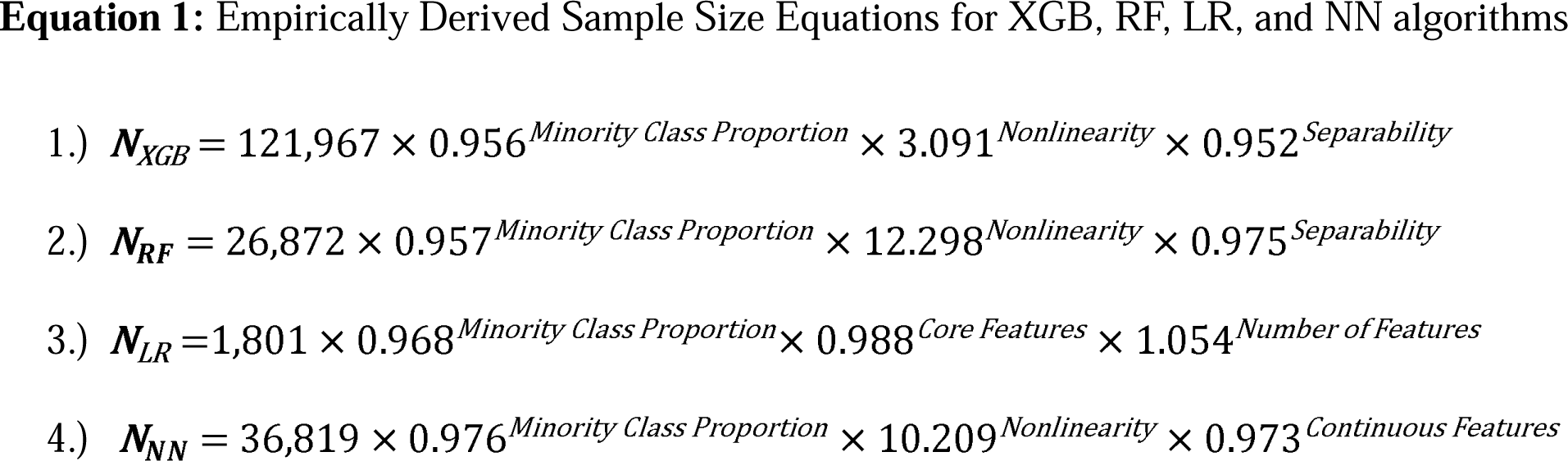

**Table 5:** Multivariable Negative Binomial Regression - Data-Specific Characteristics Effect on Predicted Sample Size

| Variable | XGB | RF | LR | NN |
| --- | --- | --- | --- | --- |
| Intercept | 121,967 [57,883-262,108] | 26,872 [8,118-92,378] | 1,801 [904-3,809] | 36,819 [18,691-76,970] |
| Minority Class Proportion | 0.956 [0.946-0.967],<br>**$p<0.0001$** | 0.957 [0.940-0.975],<br>**$p<0.0001$** | 0.968 [0.957-0.979],<br>**$p<0.0001$** | 0.976 [0.957-0.996],<br>**$p=0.016$** |
| Separability | 0.952 [0.934-0.970],<br>**$p<0.0001$** | 0.975 [0.947-1.004],<br>$p=0.074$ | -- | -- |
| Number of Features | -- | -- | 1.054 [1.034-1.076],<br>**$p<0.0001$** | -- |
| Continuous Features (%) | -- | -- | -- | 0.973 [0.950-0.997],<br>**$p=0.020$** |
| Core Linear Features (%) | -- | -- | 0.988 [0.980-0.996],<br>**$p=0.005$** | -- |
| Dataset Nonlinearity | 3.091 [2.011-4.922],<br>**$p<0.0001$** | 12.298 [5.826-28.791],<br>**$p<0.0001$** | -- | 10.209 [4.274-26.569],<br>**$p<0.0001$** |
| **Metrics** |  |  |  |  |
| Adj. Psuedo-R<sup>2</sup> | 0.845 | 0.808 | 0.798 | 0.665 |

For XGB and RF, minority class proportion, separability, and nonlinearity were the top three variables selected. For logistic regression, minority class proportion, number of features, and percentage of core features were the top three variables. For neural networks, the top three variables were minority class proportion, number of features, and nonlinearity. The direction and magnitude of coefficient estimates from multivariable models were similar to those obtained from univariable models (Table 5). Deviance-based R^2^ statistics, adjusted for the number of predictors added, were 0.845 (XGB), 0.808 (RF), 0.798 (LR), and 0.665 (NN) (Table 5). This indicated that the dataset-specific predictors explained a majority (66.5% - 84.5%) of the total deviance in the data among all four models, although the neural network model was weaker than the other three. Figure 4 shows the predicted sample sizes estimated from each algorithm-specific model at a variety of levels for each predictor. As can be seen, for all four classification algorithms, a balanced class ratio (50% cases versus 50% controls) resulted in the lowest predicted sample sizes.

**Figure 4:**
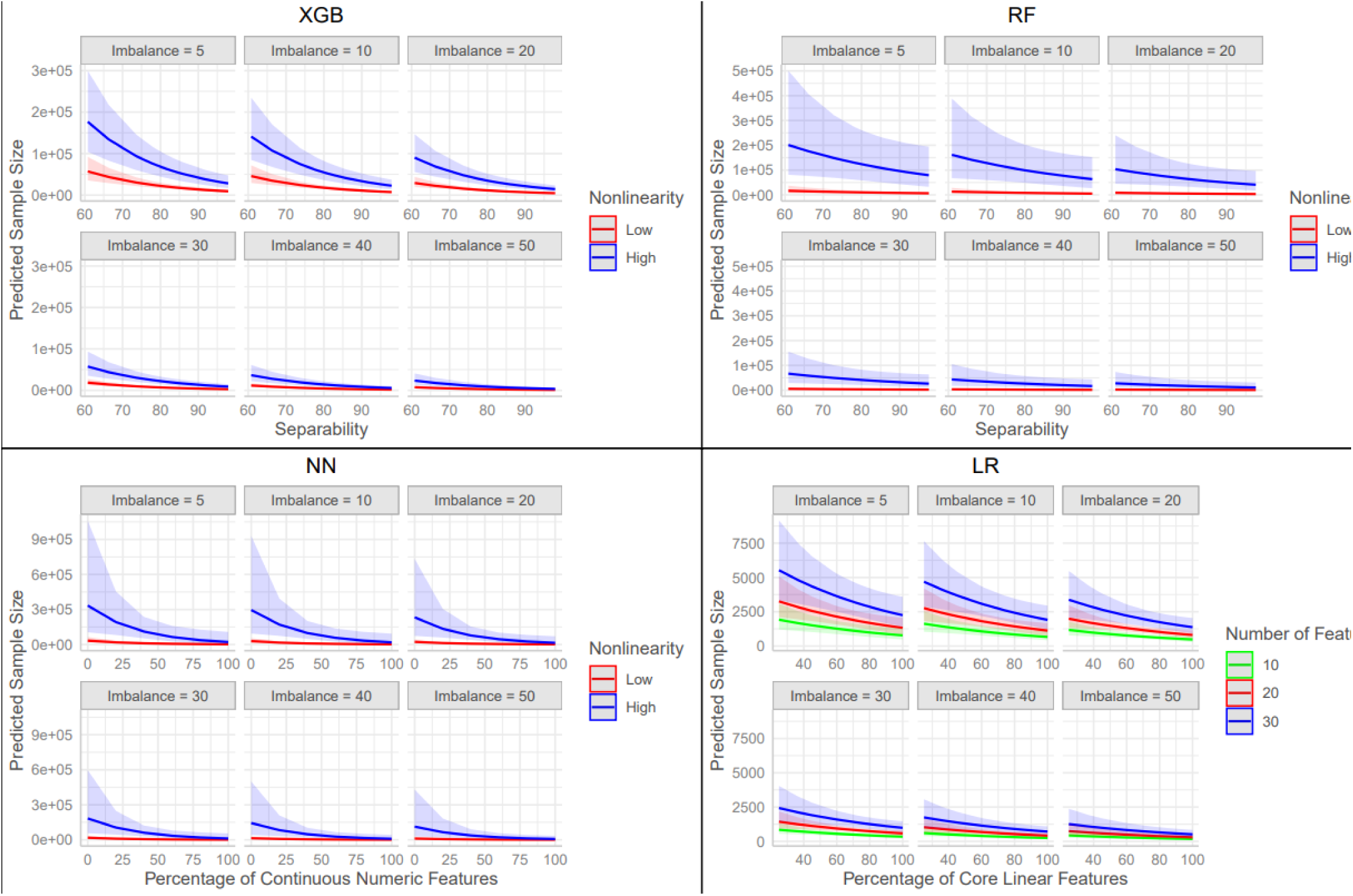
Fitted values derived from the four final negative binomial regression models for each classification algorithm. Shaded lines represent 95% confidence intervals. Imbalance = minority class proportion × 100. Separability = Full-dataset AUC × 100. Nonlinearity “low” : < 5%, “high” : ≥ 5%.

### Expected Performance at Pre-Specified Sample Size

From the empirical learning curve data, four additional models were built, examining the expected performance at each sample size point. The supplementary section includes a visualization of the AUC difference (between the full-dataset AUC and the AUC calculated at each *n*) for all datasets and all four algorithms. When values from the primary analysis (see: Table 3) were plugged into these models, we expected that the average output would be 2%, by definition. We found that all models provided predictions near this value (XGB: 1.84% on average, RF: 1.96%, LR: 1.78%, NN: 2.61%), and only the NN model tended to underestimate performance by more than a 0.5% difference in AUC.

## Discussion

In this study, we performed a learning curve analysis of sixteen datasets over four different classification algorithms. From this, we identified the expected samples needed to reach AUCs within two percentage points of those measured in the full dataset. We then examined the effects of dataset-specific characteristics on expected sample sizes, and provided formulas that can be used to predict the necessary sample size in a new dataset. We found that logistic regression required the smallest sample size (median *n* = 696) but performed slightly worse, on average, compared to more complicated algorithms. Random forest (median *n* = 3,404) and XGBoost algorithms (median *n* = 9,960) required larger sample sizes, as expected. Neural networks required the largest sample size (median *n* = 12,998) and also had the most variability over the sixteen datasets. Dataset-specific variables that altered expected sample sizes varied by algorithm, but the class imbalance of the outcome, the strength and number of features, and nonlinearity of the predictors were among the most influential characteristics.

This study provides a simple framework for determining sample size in the context of four popular machine learning algorithms. Most of the dataset-specific characteristics that we examined can be reasonably guessed or influenced before the study begins. For example, researchers can examine prior studies in their field of interest to determine a reasonable range for separability. For minority class proportion, which was a key selected feature for all four models, we observed that an optimal class balance (50% cases, 50% controls) led to the lowest predicted sample sizes, with each additional percentage point of balance decreasing the needed *n* by a multiplier of 0.96-0.98.

Additionally, researchers can use feature engineering to control the quality and overall number of predictors included in their models. As we have determined in this study, a smaller number of strong predictors will generally require less sample size than a large and noisy predictor set, supporting the idea that more features is not always ideal [32]. Dataset nonlinearity is less intuitive to guess prior to data collection. In general, we found that datasets with nonlinearity values of at least 5.0% required approximately 3-12 times the amount of sample to reach convergence, depending on algorithm. However, in this study, 13/16 (81.3%) of the datasets had values under 5.0%, which means that high values of dataset nonlinearity may be uncommon.

Again, prior studies where both simple (LR) and complex (NN/RF/XGB) methods are compared can help researchers determine if this value will be high. As a last resort, researchers can simply calculate expected sample sizes for both scenarios (< 5.0% and ≥ 5.0%) using the model equations presented in this study and discuss the implications. It is also important to note that the effect of nonlinearity (and other dataset-specific characteristics) on estimated sample sizes is diminished when the class imbalance is optimized. This is due to the multiplicative nature of the negative-binomial regression models and is illustrated in Figure 4. Thus, it is critical that researchers aim to collect a sample with the most balance between cases and controls.

As a secondary aim, we built models that inverted the learning curve data from the primary analysis, enabling prediction of the expected performance gap at some user-defined sample size. Because the same set of dataset-specific characteristics were used as covariates in these models, researchers can enter their expected dataset-specific characteristics and a sample size (or range of sample sizes), and will be given a prediction for the expected AUC difference at that *n*. Because we already derived the *n* where the AUC was 2 percentage points less than the full-dataset AUCs from our primary analysis, we were able to test the internal validity by plugging in these predicted sample sizes and seeing how close to 2% the mean output of each model was. We found that, on average, the XGB, RF, and LR models were quite close to this value, while the NN model tended to underpredict performance slightly more. Regardless, some error was expected due to variability in the empirical learning curve data. These models can provide valuable insight on the expected performance of each algorithm at a range of sample sizes, and may guide the choice of classification method, if researchers are limited by data collection.

To our knowledge, no prior study has presented specific formulas for calculating sample size within the context of machine learning for a binary outcome in the field of healthcare/clinical data analysis. We examined sixteen datasets, which is a relatively large amount in this area of research - similar learning curve analyses have typically examined less than ten [14,15].

Assessment of more, potentially non-open access datasets, could strengthen these models and provide clearer insight on dataset-specific effects on expected sample size, although the fact that we still observed many statistically significant relationships even with an effective sample size of *n=*16 is a strength of the study. Additionally, the datasets we examined had relatively low (< 50) numbers of features – generalization of the formulas presented in this study may not extrapolate to datasets with larger amounts of features. Finally, it is important to note that machine learning, specifically algorithms like random forest and XGBoost, can still outperform traditional parametric methods even if the sample size is limited (i.e. under *n* = 5,000), and when hyperparameter tuning is implemented [33, 34, 35]. Therefore, these guidelines should serve as a supplement, giving a general idea of how much sample is expected to reach a point of “diminishing returns,” where large amounts of additional data will only increase the AUC marginally.

Future research in this area could examine different outcome types, such as multi-class or survival endpoints. Additionally, a more in-depth examination of XGBoost/RF/NN hyperparameters would be impactful, as all of the equations developed in this study considered only the default hyperparameter values. Finally, stacked machine learning methods [36], or different gradient-boosted tree algorithms such as CatBoost [37] or LightGBM [38] could be investigated.

## Supporting information

Supplementary Section

## Data Availability

All data is publicly available online. Exact locations can be found in the Supplementary Section.

## Data Availability and Access

All datasets in CSV format and R Code will be uploaded to GitHub following publication.

An RShiny app will become available for public use following publication, where users can calculate expected sample size from the equations and models generated in this study.

