## Supplementary Section for "Empirical Sample Size Determination for Popular Classification Algorithms in Clinical Research"

**Supplementary Table #1 :** Learning Curve Details

*In order : number of points examined, range of potential sample sizes.

| **Dataset** | **XGB*** | **RF*** | **LR*** | **NN*** | **Curve Fitting Method**** |
| --- | --- | --- | --- | --- | --- |
| Cardio | 10, [500-50,000] | 10, [500-50,000] | 25, [100-5,000] | 12, [500-50,000] | 1,1,2,2 |
| Diabetes130 | 10, [500-50,000] | 10, [500-50,000] | 25, [100-5,000] | 12, [500-50,000] | 1,1,3,3 |
| NoShow | 10, [500-50,000] | 10, [500-50,000] | 24, [296-5,000] | 12, [500-50,000] | 3,1,3,3 |
| BreastTumor | 10, [500-50,000] | 10, [500-50,000] | 25, [100-5,000] | 12, [500-50,000] | 3,3,2,2 |
| Diabetes | 10, [500-50,000] | 10, [500-50,000] | 25, [500-10,000] | 12, [500-50,000] | 1,1,2,2 |
| COVID-19 | 10, [500-50,000] | 10, [500-50,000] | 25, [100-5,000] | 12, [500-50,000] | 1,1,2,1 |
| LOS | 10, [500-50,000] | 10, [500-50,000] | 25, [100-5,000] | 12, [500-50,000] | 3,3,2,2 |
| CDC Heart Disease (2020) | 10, [500-50,000] | 10, [500-50,000] | 25, [100-10,000] | 12, [500-50,000] | 1,3,2,2 |
| CDC Heart Disease (2022) | 10, [500-50,000] | 10, [500-50,000] | 25, [100-10,000] | 12, [500-50,000] | 3,3,2,2 |
| Heart | 10, [500-50,000] | 10, [500-50,000] | 25, [100-5,000] | 12, [500-50,000] | 1,3,2,1 |
| Hepatitis | 10, [500-50,000] | 10, [500-50,000] | 25, [100-5,000] | 12, [500-50,000] | 1,1,2,2 |
| Lymph | 10, [500-50,000] | 10, [500-50,000] | 25, [100-5,000] | 12, [500-50,000] | 1,3,2,1 |
| Pharynx | 10, [500-50,000] | 10, [500-50,000] | 25, [100-5,000] | 12, [500-50,000] | 3,1,2,1 |
| Cholesterol | 10, [500-100,000] | 10, [500-200,000] | 25, [100-5,000] | 11, [500-250,000] | 3,1,3,3 |
| Dermatology | 10, [500-50,000] | 10, [500-50,000] | 25, [500-10,000] | 12, [500-50,000] | 1,1,2,3 |
| PBC | 10, [500-50,000] | 10, [500-100,000] | 25, [100-5,000] | 11, [500-100,000] | 3,3,2,3 |

**In Order : XGB, RF, LR, NN. 1 = Power Law (*c* estimated), 2 = Power Law (*c* fixed to full-dataset AUC), 3 = log-linear regression.

**Supplementary** : Dataset Description and Processing Steps :

**Cardio :** Obtained from OpenML (https://www.openml.org/search?type=data&status=active&id=45547).

Features included were age (continuous numeric), gender (binary), height (continuous numeric), weight (continuous numeric), SBP (continuous numeric), DBP (continuous numeric), chol (ordinal), glucose (ordinal), smoking (binary), alcohol (binary), PhysicalActivity (binary), and CVD (outcome). No further processing steps were considered.

**Diabetes130 :** Obtained from OpenML (<https://www.openml.org/search?type=data&status=active&sort=qualities.NumberOfInstances&id=4541>).

Values with “?” were changed to NA. NA values were imputed using na.roughfix, from the RandomForest R package.

Features included were race (Caucasian = 1, Other = 0, binary), gender (Male = 1, Other = 0, binary), age (Under 20 = 0, Over 20 = 1, binary), time in hospital (continuous numeric), num_lab_procedures (continuous numeric), num_procedures (discrete numeric), num_medications (continuous numeric), number_outpatient (continuous numeric), number_emergency (continuous numeric), number_inpatient(continuous numeric), number_diagnoses (continuous numeric), metformin (Steady = 1, No = 0, binary), repaglinide (Steady = 1, No = 0, binary), nateglinide (Steady = 1, No = 0, binary), chlorpropramide (Steady = 1, No = 0, binary), glimepiride (Steady = 1, No = 0, binary), acetohexamide (Steady = 1, No = 0, binary), glipizide (Steady = 1, No = 0, binary), glyburide (Steady = 1, No = 0, binary), tolbutamide (Steady = 1, No = 0, binary), pioglitazone (Steady = 1, No = 0, binary), rosiglitazone (Steady = 1, No = 0, binary), acarbose (Steady = 1, No = 0, binary), miglitol (Steady = 1, No = 0, binary), troglitazone (Steady = 1, No = 0, binary), tolazamide (Steady = 1, No = 0, binary), citoglipton (Steady = 1, No = 0, binary), insulin (Steady = 0, Other = 1, binary), glyburide.metformin (Steady = 1, No = 0, binary), glipizide.metformin (Steady = 1, No = 0, binary), glimepiride.metformin (Steady = 1, No = 0, binary), metformin.rosiglitazone (Steady = 1, No = 0, binary), metformin.pioglitazone (Steady = 1, No = 0, binary), change (Ch = 1, Other = 0, binary), diabetesMed (Yes = 1, Other = 0, binary), readmitted (No = 0, Other = 1, outcome).

**NoShow :** Obtained from OpenML (<https://www.openml.org/search?type=data&status=active&sort=qualities.NumberOfInstances&order=desc&id=43439>).

Features included were gender (Male = 1, F = 0, binary), Age (continuous numeric), Scholarship (yes = 1, no = 0, binary), Hypertension (yes = 1, no = 0, binary), Diabetes (yes = 1, no = 0, binary), Alcoholism (yes = 1, no = 0, binary), Handicap (yes = 1, no = 0, binary), SMS (yes = 1, no = 0, binary), NoShow (1 = yes, 0 = no, outcome). No further processing steps were considered.

**BreastTumor** : Obtained from OpenML (<https://www.openml.org/search?type=data&status=active&id=1201>)

Features included were age (continuous numeric), menopause (1=premenopausal, 0=other, binary), inv.nodes (continuous numeric), node.capes(1 = yes, 0 = no, binary), deg.malig(ordinal), breast(1 = yes, 0 = no, binary), breast.quad (1 = central, 0 = other, binary), irradiation (1 = yes, 0 = no, binary), class (continuous numeric), recurrence (r = 1, other = 0, outcome). No further processing steps were considered.

**Diabetes** : Obtained from UCI ML Repository (<https://archive.ics.uci.edu/dataset/891/cdc+diabetes+health+indicators>)

Features included were HighBP (1 = yes, 0 = no, binary), HighChol (1 = yes, 0 = no, binary), CholCheck (1 = yes, 0 = no, binary), BMI (continuous numeric), Smoker (1 = yes, 0 = no, binary), Stroke (1 = yes, 0 = no, binary), HeartDiseaseOrAttack (1 = yes, 0 = no, binary), PhysActivity (1 = yes, 0 = no, binary), Fruits (1 = yes, 0 = no, binary), Veggies (1 = yes, 0 = no, binary), HvyAlcoholConsump (1 = yes, 0 = no, binary), AnyHealthCare (1 = yes, 0 = no, binary), NoDocbcCost (1 = yes, 0 = no, binary)¸GenHlth (Ordinal), MentHlth (continuous numeric), PhysHlth (continuous numeric), DiffWalk (1 = yes, 0 = no, binary), Sex (1 = Male, 0 = Female, binary), Age (continuous numeric), Education (Ordinal), Income (Ordinal), Diabetes_binary (1 = yes, 0 = no, binary). No further processing steps were considered.

**COVID-19** : Obtained from OpenML (<https://www.openml.org/search?type=data&status=any&id=43428>)

Values with “99” or “97” were changed to NA. NA values were imputed using na.roughfix, from the RandomForest R package.

Features included were : SEXO (1 = M, 0 = F, binary), TIPO_PACIENTE (1/0, binary), NEUMONIA (1 = yes, 0 = no, binary), EDAD (continuous numeric), NACIONALIDAD (0/1, binary), HABLA_LENGUA_INDIG (1 = yes, 0 = no, binary), DIABETES (1 = yes, 0 = no, binary), EPOC (1 = yes, 0 = no, binary)¸ASMA (1 = yes, 0 = no, binary), INMUSUPR (1 = yes, 0 = no, binary), HIPERTENSION (1 = yes, 0 = no, binary), OTRA_COM (1 = yes, 0 = no, binary), CARDIOVASCULAR (1 = yes, 0 = no, binary), OBESIDAD (1 = yes, 0 = no, binary), RENAL_CRONICA (1 = yes, 0 = no, binary)¸TABAQUISMO (1 = yes, 0 = no, binary), RESULTADO (1 = 0, 2= 1, outcome). No further processing steps were considered.

**LOS** : Obtained from OpenML (<https://www.openml.org/search?type=data&status=active&id=43550>)

Values with “?” were changed to NA. NA values were imputed using na.roughfix, from the RandomForest R package.

Features included were HospitalType (1 = 1, 0 = other, binary), availableextrarooms (continuous numeric), department (gynecology = 0, other = 1, binary), WardType (R = 0, Other = 1, binary), WardFacility (F = 0, other = 1, binary), BedGrade (2 = 0, Other = 1, binary), TypeOfAdmission (Trauma = 0, Other = 1, binary), Severity (Extreme = 1, Other = 0, binary), Visitors (continuous numeric), Age (81 and older = 1, Other = 0, binary), Deposit (continuous numeric), LOS (>100 days = 1, Less than 100 days = 0, outcome).

No further processing steps were considered.

**CDC Heart Disease (2020)** : Obtained from Kaggle (<https://www.kaggle.com/datasets/kamilpytlak/personal-key-indicators-of-heart-disease>)

Features included were BMI (continuous numeric), Smoking (1 = yes, 0 = no, binary), AlcoholDrinking (1 = yes, 0 = no, binary)¸Stroke (1 = yes, 0 = no, binary), PhysicalHealth (continuous numeric), MentalHealth (continuous numeric), DiffWalking (1 = yes, 0 = no, binary), Sex (1 = MALE, 0 = Female, binary), AgeCategory (1 = 50+, 0 = under 50, binary), Race (White = 1, Other = 0, binary), Diabetic (1 = yes, 0 = no, binary), PhysicalActivity (1 = yes, 0 = no, binary), GenHealth (Good, very good, excellent = 1, other = 0, binary), SleepTime (continuous numeric), Asthma (1 = yes, 0 = no, binary), KidneyDisease (1 = yes, 0 = no, binary), SkinCancer (1 = yes, 0 = no, binary), HeartDisease (1 = yes, 0 = no, outcome).

No further processing steps were considered.

**CDC Heart Disease (2022)** : Obtained from Kaggle (<https://www.kaggle.com/datasets/kamilpytlak/personal-key-indicators-of-heart-disease>)

NA values were imputed using na.roughfix, from the RandomForest R package.

Features included were State (“Connecticut", "Delaware", "District of Columbia", "Maine", "Maryland", "Massachusetts", "New Hampshire", "New Jersey", "New York", "Pennsylvania", "Rhode Island", "Virginia", "Vermont" = 1, Other = 0, binary), GeneralHealth (0 = Poor, 1 = Other, binary), PhysicalHealthDays (continuous numeric), MentalHealthDays (continuous numeric), LastCheckupTime (within past year = 1, other = 0, binary), PhysicalActivities (1 = yes, 0 = no, binary), SleepHours (continuous numeric), RemovedTeeth (1 = other, 0 = none of them, binary), HadHeartAttack (1 = yes, 0 = no, binary), HadAngina (1 = yes, 0 = no, binary), HadStroke (1 = yes, 0 = no, binary), HadAthsma (1 = yes, 0 = no, binary), HadSkinCancer (1 = yes, 0 = no, binary), hadCOPD (1 = yes, 0 = no, binary), HadDepressiveDisorder (1 = yes, 0 = no, binary)¸HadKidneyDisease (1 = yes, 0 = no, binary)¸HadArthritis (1 = yes, 0 = no, binary), HadDiabetes (1 = yes, 0 = no, binary), DeafOrHardOfHearing (1 = yes, 0 = no, binary)¸BlindOrVisionDifficulty (1 = yes, 0 = no, binary)¸DifficultyConcentrating (1 = yes, 0 = no, binary), DifficultyWalking (1 = yes, 0 = no, binary)¸DifficultyDressingBathing (1 = yes, 0 = no, binary), DifficultyErrands (1 = yes, 0 = no, binary), SmokerStatus (never smoked = 0, other = 1, binary), eCiggaretteUsage (0 = never, 1 = other, binary), ChestScan (1 = yes, 0 = no, binary), RaceEthnicityCategory (0 = non-hispanic white, 1 = other, binary), AgeCategory (over 65 = 1, other = 0, binary), HeightInMeters (continuous numeric), WeightInKilograms (continuous numeric), BMI (continuous numeric), Alcohol Drinkers (1 = yes, 0 = no, binary), HIVTesting (1 = yes, 0 = no, binary)¸FluVaxLast12 (1 = yes, 0 = no, binary), PneumoVaxEver (1 = yes, 0 = no, binary)¸TetanusLast10Tdap (yes, received tdap = 1, other = 0, binary), CovidPOS (1 = yes, 0 = no, binary)¸HighRiskLastYear (1 = yes, 0 = no, outcome). No further processing steps were considered.

**Heart** : Obtained from OpenML (<https://www.openml.org/search?type=data&status=active&sort=qualities.NumberOfInstances&id=267>)

Features included were age (continuous numeric), sex (0/1, binary), chest (continuous numeric), resting_blood_pressure (continuous numeric), serum_cholesterol (continuous numeric), fasting_blood_sugar (0/1, binary), resting_electrocardiographic_results (ordinal), maximum_heart_rate_achieved (continuous numeric), exercise_induced_angina (0/1, binary), oldpeak (continuous numeric), slope (ordinal), number_of_major_vessels (ordinal), thal (ordinal), class (present = 1, other = 0, outcome). No further processing steps were considered.

**Hepatitis** : Obtained from OpenML (<https://www.openml.org/search?type=data&status=active&sort=qualities.NumberOfInstances&id=269>)

Features considered were AGE (continuous numeric), SEX (1 = male, 0 = female, binary), STEROID (1 = yes, 0 = no, binary), ANTIVIRALS (1 = yes, 0 = no, binary), FATIGUE (1 = yes, 0 = no, binary)¸MALAISE (1 = yes, 0 = no, binary)¸ANOREXIA (1 = yes, 0 = no, binary), LIVER_BIG (1 = yes, 0 = no, binary), LIVER_FIRM (1 = yes, 0 = no, binary)¸SPLEEN_PALPABLE (1 = yes, 0 = no, binary)¸SPIDERS (1 = yes, 0 = no, binary)¸ASCITES (1 = yes, 0 = no, binary)¸VARICES (1 = yes, 0 = no, binary), BILIRUBIN (continuous numeric), ALK_PHOSPHATE (continuous numeric), SGOT (continuous numeric), ALBUMIN (continuous numeric), PROTIME (continuous numeric), HISTOLOGY (1 = yes, 0 = no, binary), Class (0 = LIVE, 1 = other, outcome). No further processing steps were considered.

**Lymph** : Obtained from OpenML (<https://www.openml.org/search?type=data&status=active&sort=qualities.NumberOfInstances&id=249>)

Features considered were lymphatics (0 = normal, 1 = other, binary), block_of_affere (0 = no, 1 = yes, binary), bl_of_lymph_c (0 = no, 1 = yes, binary), By_pass (0 = no, 1 = yes, binary), extravastes (0 = no, 1 = yes, binary)¸regeneration_of (0 = no, 1 = yes, binary)¸early_uptake_in (0 = no, 1 = yes, binary), lym_nodes_dimin (0 = no, 1 = yes, binary), lym_nodes_enlar (0 = no, 1 = yes, binary), changes_in_lym (0 = round, 1 = other, binary), defect_in_node (0 = no, 1 = yes, binary)¸changes_in_stru (0 = no, 1 = yes, binary), changes_in_node (0 = no, 1 = yes, binary), special_forms (0 = no, 1 = yes, binary), dislocation_of (0 = no, 1 = yes, binary), exclusion_of_no (0 = no, 1 = yes, binary), no_of_nodes_in (continuous numeric), class (1 = metastases, 0 = other, outcome). No further processing steps were considered.

**Pharynx :** Obtained from OpenML (<https://www.openml.org/search?type=data&status=active&id=1196>)

Features included were Inst (Ordinal), sex (1 = 0, 2 = 1, binary), Treatment (2 = 1, 1 = 0, binary), Grade (1 = 0, other = 1, binary), Condition (1,0 = 0, other = 1, binary), Site (4 = 0, other = 1, binary), T (1, 2 = 0, other = 1, binary), N (0, 1 = 0, other = 1, binary), Age (continuous numeric), Entry (continuous numeric), Status (0/1 binary), class (> 750 = 1, otherwise = 0, outcome). No further processing steps were considered.

**Cholesterol** : Obtained from OpenML (<https://www.openml.org/search?type=data&status=active&sort=qualities.NumberOfInstances&id=1194>)

Features included were age (continuous numeric), sex (0/1 binary), cp (ordinal), trestbps (continuous numeric), fbs (0/1 binary), restecg (ordinal), thalach (continuous numeric), exang (0/1 binary), oldpeak (continuous numeric), slope (ordinal), ca (ordinal), thal (ordinal), num (ordinal), chol (> 200 = 0, otherwise = 1, outcome). No further processing steps were considered.

**Dermatology :** Obtained from OpenML (<https://www.openml.org/search?type=data&status=active&id=263>)

Features included were erythema (ordinal), scaling (ordinal)¸definite_borders (ordinal)¸itching (ordinal)¸koebner_phenomenon (ordinal), polygonal_papules (ordinal), follicular_papules (ordinal), oral_mucosal_involvement (ordinal), knee_and_elbow_involvement (ordinal)¸scalp_involvement (ordinal)¸melanin_incontenence (ordinal), eosinophils_in_the_inflitrate (ordinal), PNL_infiltrate (ordinal)¸fibrosis_of_the_papillary_dermis (ordinal), exocytosis (ordinal)¸acanthosis (ordinal)¸hyperkeratosis (ordinal)¸parakeratosis (ordinal)¸clubbing_of_the_rete_ridges (ordinal)¸elongation_of_the_rete_ridges (ordinal)¸thinning_of_the_suprapapillary_epidermis (ordinal), spongiform_pustule (ordinal)¸munro_microabsess (ordinal), focal_hypergranulosis (ordinal)¸disappearance_of_the_granular_layer (ordinal), vascolisation_and_damage_of_basal_layer (ordinal), spongiosis (ordinal)¸saw.tooth_appearance_of_retes (ordinal)¸follicular_horn_plug (ordinal)¸perifollicular_parakeratosis (ordinal), inflammatory_mononuclear_inflitrate (ordinal)¸band.like_infiltrate (ordinal)¸Age (continuous numeric), family_history (1 = yes, 0 = no, outcome). No further processing steps were considered.

**PBC** : Obtained from OpenML (<https://www.openml.org/search?type=data&status=active&id=1191>)

Features included were D (0/1 binary), Z1 (ordinal), Z2 (continuous numeric), Z3 (0/1 binary), Z4 (0/1 binary), Z5 (0/1 binary), Z6 (0/1 binary), Z7 (ordinal), Z8 (continuous numeric), Z9 (continuous numeric), Z10 (continuous numeric), Z11 (continuous numeric), Z12 (continuous numeric), Z13 (continuous numeric), Z14 (continuous numeric), Z15 (continuous numeric), Z16 (continuous numeric), Z17 (ordinal), class (> 3000 = 1, otherwise = 0, outcome). No further processing steps were considered.

**Supplementary Figure #1** : Empirical AUC Differences at Sample Size *N*

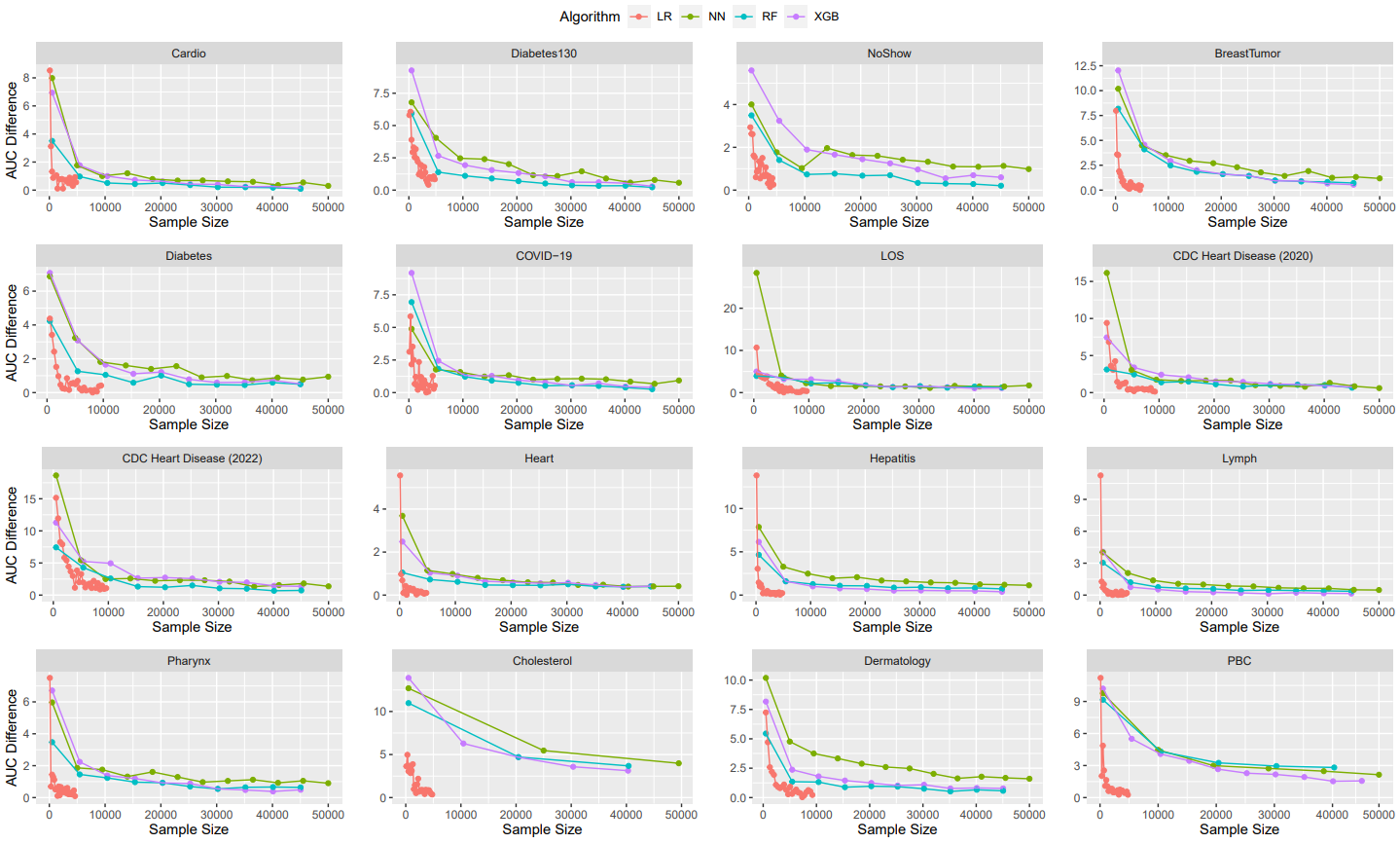
